# HAEMOPARASITIC INFECTION IN RED SOKOTO GOATS IN MAKURDI, BENUE STATE

**DOI:** 10.1101/2025.10.08.25337525

**Authors:** Shiaondo Isaac Kaha, Jighjigh Friday Gudu, David Tyover Kajo, Tavershima T Dajo

**Affiliations:** Department of Biological Sciences, Benue State University, Makurdi, Nigeria

## Abstract

Among the various factors that cause decrease in productivity and maximum yield of livestock in Sub-tropical Africa, disease is the most prominent agent. Blood samples were collected from 240 Red Sokoto breed of goats from four abattoirs (Wurukum, Wadata, North bank, and Modern Market abattoirs) in Makurdi town, between October 2017 and January 2018, to check for the presence of the parasites. Of the 240 samples examined, 59 were positive for parasites,14(5.8%) of the positive samples were male and 45(18.8%) were female. Wurukum and Wadata markets had a tie of 19(7.9%) infected, North Bank, 14(5.8%) and Modern market, 7(2.9%). Parasites seen were *Babesia spp* [33(13.8%)], *Anaplasma spp* [13(5.4%)], *Theileria spp* [9(3.8%)]and *Trypanosoma spp* [4(1.7%)]. Cases of multiple infections were also recorded in four (4) goats. *Babesia spp* and *Anaplasma spp* topped chart with two (2) animals positive, while *Babesia spp/Theileria spp* and *Theileria spp/Anaplasma spp* both occurred in one animal each. This study therefore confirms the presence of these parasites in goats slaughtered at the various abattoirs in Makurdi and therefore paves way for in-depth studies on the conditions under which these animals are reared, their relative productivity and brooding capabilities with other animals kept under healthier and disease-controlled environment. Also, their conditions before they are brought to the slaughter house as confirmatory tests and hence, a measure to help tackle their spread and further destruction which they cause to the livestock system of the country.

## 1. Introduction

Sheep and goats form an important part of livestock industry in the sub- Saharan Africa. They serve as valuable supplement to cattle in terms of animal protein supply for the teaming population including provision of manure for field crops. It has been established that over 90% of sheep and goats in the sub- Saharan Africa are found in East and West Africa (Jatau *et al*., 2011). These animals are important source of investment especially in the rural areas including Nigeria, where livestock is regarded as capital investment in the absence of banking facilities as well as serving as an important source of meat, milk, skin and socio-cultural values. Traditionally, the nomadic herdsmen (pastoralists),also keep sheep and goats which graze along with cattle. This practice seems to maximize economic benefits owing to the small ruminants high fertility and early maturity as well as their adaptability to the environment (Ademosun., 1988).

The benefits derived from sheep and goats in the tropics are far below the expected due mainly to low productivity. This is due to numerous factors of which disease is the most important (Akerejola *et al*., 1979). Goats in Sub-Sahara Africa may be infected with a wide variety of parasites among which the gastrointestinal parasitic infection are the most commonest and these include Helminthic infections especially *Haemonchus contortus*, Trichostrongilus, *Cooperia* and protozoan diseases including Coccidiosis (Ngole *et al*., 2001,Okaiyeto*et al*., 2008) as well as economically important Vector-borne prokaryotic and eukaryotic haemoparasites such as the *Rickettsiae, Anaplasma* and *Ehrlichia* (cowdria) and the protozoan parasites *Theileria, Babesia* and *Trypanosoma* (Bell-Sakyi *et al*., 2004, Okaiyeto *et al*., 2008)

Haemoprotozoan parasites are the main livestock production constraints all over the world, causing serious economic losses, tick and tick-borne diseases(T and TBDs) still remain a major threat to animals in tropical and sub-tropical countries including Nigeria. In case of these blood parasites infection, up to 75% erythrocytes may be destroyed in fatal cases and even in milder infection, so many erythrocytes are destroyed, then a severe aneamia results. Babesiosis, Theileriosis and Ehrelichosis (cowdriosis and Anaplasmosis) are the major TBDs that cause serious diseases among central and East African animals including goats. This is however not to completely neglect the effect and subsequent damage caused by *Trypanosoma spp* which causes sleeping sickness to animals.

This study was undertaken to know the ubiquity of haemoparasites prevalent in goats in Makurdi abattoirs, Benue State. This will further pave way for launching sustainable animal disease controlling/minimizing in so many parts of the country (Nigeria) and of course, the whole world.

The parasites of livestock find the tropical environment very favorable for their growth and development.(Panye, 1990) The direct losses caused by the parasites are attributed to acute illness and death, premature slaughter and rejection of some body parts at meat inspection.

Indirect losses include the reduction of productivity potential such as decreased growth rate, weight loss in young growing animals and late maturity of slaughter stock (Hansen and Perry, 1994).

Proper understanding of the epidemiology of disease causing agents is a pre requisite for the rational design of effective preventive and the control program against the disease. Although studies have been carried out with respect to epidemiology of blood parasites hence the need for the extension of such studies to small ruminants. This study is therefore targeted at providing relevant information in this regard.

## 2. Materials and Method

### 2.1 Study Area

The study was carried out in Makurdi Local Government area of Benue State. Makurdi Local Government area was founded in 1976. It lies on the South Bank of the Benue River. Makurdi rapidly developed into a market and transportation center. Following the division of Benue Plateau states into two states in 1976, Makurdi was selected as the capital of Benue state.

Council wards in Makurdi Local Government Area include Agan, Mbalagh, North Bank, Central South Mission, Ankpa/Wadata, Modern Market, Fiidi, Bar and Wailomayo council wards. Major indigenous groups in the local government Area are Tiv, Idoma, Igede, Jukum and Etulo. Others are Hausa, Igbo and Yoruba. while farming is the major occupation in the local government, education is the dominant industry (Makurdi LGC, 2011).

Makurdi local government is also endowed with the establishment of many educational institutions, prominent among which are the Murtala College of Arts, Science and Technology (1976), the federal university of technology, an Assemblies of God Commercial Institute, a Government Craft School, the federal University of Agriculture and Benue State University. Other institutions like Nursery/primary and secondary schools abound. Some of the schools are owned by the government, mission, non-governmental organizations (NGOs) and private individuals. (Makurdi LGC, 2011) Makurdi has a population of about 297,398 people (NPC, 2006). Makurdi has majorly six (6) abattoirs; Wurukum Market abattoir, Wadata Market Abattoir, Railway Market Abattoir North bank Market abattoir, Modern market abattoir, and High Level Market abattoir.

This study made the choice of all these abattoirs for the sake of diversity in species availability in all these abattoirs so as to check the distribution of these parasites in the abattoirs and from the various places of production of the goat species.

### 2.2 Sampling method

Blood samples were collected from slaughtered goats randomly, at the abattoirs from November to December 2017.

A random sample of two hundred and forty (240) goats was done across the six abattoirs in Makurdi Local Government Area, forty from each. The research used simple Random Sampling technique to select the Goat. The Red Sokoto breed of Goats were used because, past researchers in the North Central axis and Nigeria to some extent, have not been able to deal specifically with this species. These species of goats are commonly reared in Benue State, and so it forms part of the meat supply for the town.

### 2.3 Sample Size Determination

Sample size was determined using the formula:

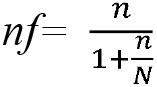

where *nf=* Sample size when the population is less than 10,000

*n*=the desired sample size when the population is more than 10,000 i.e. 240

N=the estimate of the population size i.e 297,398

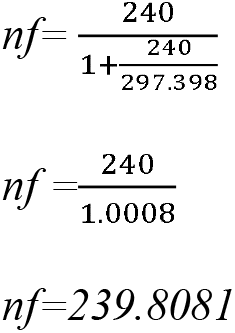

Source: Bartlett (2001)

### 2.4 Sample Collection

Five (5) mls of blood was collected from the severed jugular vein into a bijou bottle containing Ethylene Diamine Tetra Acetate (EDTA). The samples were properly labeled and transported to the laboratory immediately in ice packs.

### 2.5 Parasitological Analysis

On arrival to the laboratory, the blood samples were immediately examined for the presence of parasites using wet blood film, Giemsa stained thin blood. Wet blood films were prepared as described by Cheesbrough (1999) as follows;

A drop of blood was placed onto a clean glass slide and then followed by placing a clean cover slip on the drop of blood which spread the latter into a monolayer of cells. This was then examined for trypanosomes movement using 40X objective lens.

A thin blood smear was prepared from each blood sample by placing a drop of blood on one end of a clean glass slide, a spreader was then used to spread the blood by allowing the spreader to touch the blood and then spread gently but firmly along the surface of the horizontal slide so that the blood is dragged behind the spreader to form the film with a feathered edge. The film was air-dried and fixed with methanol for three to five minutes, stained with giemsa followed by buffer dilution and stained for twenty five to thirty minutes then rinsed with distilled water and allowed to dry. The smear was examined with 100X magnification (oil immersion) for presence of parasites and identification according to Cheesbrough (1999).

### 2.6 Statistical Analysis

SPSS was used to analyze raw data gotten from laboratory experiment results.

## 3. Results

Table 1 shows the heamoparasitic rate of infection in the sampled goats. Of the 240 goats sampled, 59 were infected with parasites. The rate of infection in male goats was 5.8% and that of female goats was 18.8%. Statistical analysis showed that there was no significant difference(p>0.05) in sex.

**Table 1:**
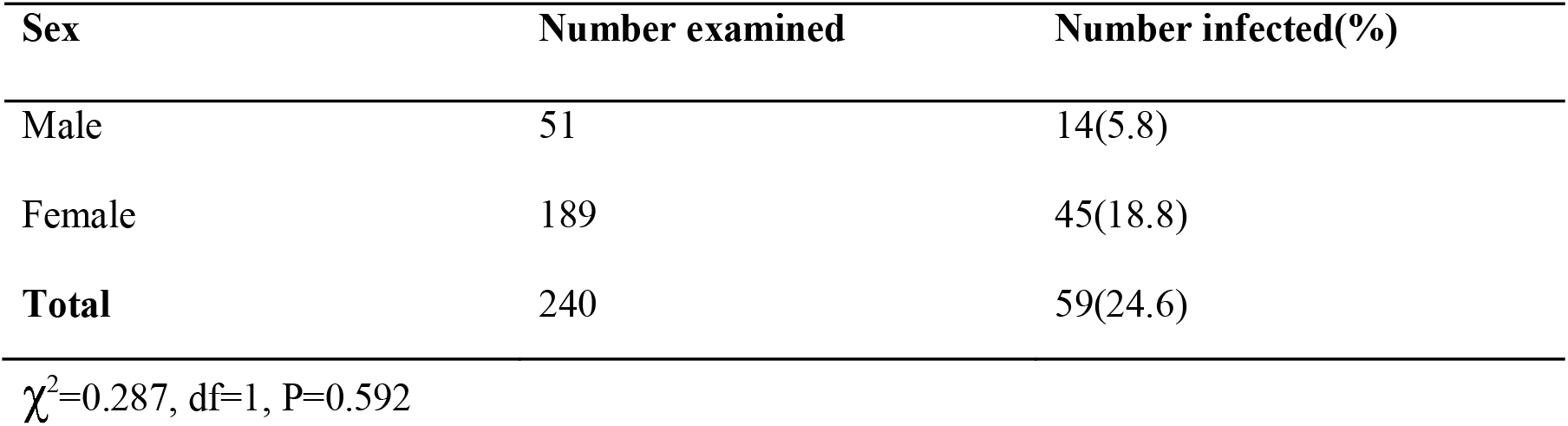
Parasitic infection in goats according to sex.

**Table 2:** Rate of infection according to the type of parasite seen.

| Parasite | Number positive | Rate(%) |
| --- | --- | --- |
| *Babesia spp* | 33 | 13.8 |
| *Anaplasma spp* | 13 | 5.4 |
| *Trypanosoma spp* | 4 | 1.7 |
| *Theileria spp* | 9 | 3.8 |
| **Total** | 59 | 24.6 |

while the least was *Trypanosoma spp* (1.7%). There was a significance between infection and parasites observed (p<0.05).

Table 3 shows that, infection inrelation to thepresence ofmore than one parasite is significant, statistically. Multiple infection involving *Anaplasma spp* and *Babesia spp* (0.8%) compared to *Babesia spp Theileria spp* and *Theileria spp/Anaplasma spp* (0.4%).

**Table 3:** Multiple Infection of Parasites in Goats.

| Multiple infection | Number positive | Rate (%) |
| --- | --- | --- |
| *Babesia spp* and *Theileria spp* | 1 | 0.4 |
| *Anaplasma spp* and *Babesia spp* | 2 | 0.8 |
| *Theileria spp* and *Anaplasma spp* | 1 | 0.4 |
| **Total** | 4 | 0.02 |

Table 4 below shows that, of the four (4) locations sampled, Wadata and Wurukum had the highest infection rate [19(7.9%)] while the least infected location was in Modern market [7 (2.9%)]. Statistical analysis showed that there was no significant difference between infection and location of the goats (p>0.05).

**Table 4:** Parasitic Infection of Goats According to Location.

| Location | Number examined | Number infected(%) |
| --- | --- | --- |
| Wurukum | 71 | 19(.9) |
| North- Bank | 61 | 14(5.8) |
| Modern Market | 43 | 7(2.9) |
| Wadata | 65 | 19(7.9) |
| **Total** | 240 | 54(24.6) |

## 4. Discussion

The species of haemoparasites that were found and reported in this study are in absolute conformity with those reported earlier by researchers on the range of haemoparasites found among livestock in Nigeria and specifically, in the North Central Region (Kamani *et al*., 2010). *Babesia spp* with occurrence rate of 13.8% and the highest among the four haemoparasites observed also agrees with the findings of (Quadeer *et al*., 2015, Kamani *et al*., 2010) as being the highest occurring parasites. The relatively small number of *Tryanosoma spp*, indicates that small ruminants play little or no role in their (Trypanosomes) epidemiology.

More so, upon investigation at the abattoirs where supply and slaughter of the goats is done, it was inferred that most supply of the goats is from the far Northern part of Nigeria which is normally not characterized by thick forest and extreme humidity and so do not support the transmission of trypanosomes by tick vectors as can be found in the rainforest regions of the country. It is also not new that camels are herded together with small ruminants in some parts of Northern Nigeria. Therefore, disease transmission among these animals is high as a result.

There was also no observed statistical significance between infection and sex of the goats and this may be due to the reason that no sex has been found to possess greater resistance to the parasites, this is not however, to undermine the fact that the number of female animals sampled were infected more with the parasites and this can be attributed to the common reason that the females are kept for longer periods than the males at their locations of breeding, mostly for the purpose of reproduction and hence are more susceptible or more open to getting in contact with the parasites due to the longer duration which they live.

The locations of sampling did not also show any significant statistical difference between each other on the basis of the number of parasites found. It is however important to note that the conditions under which the animals are kept preparatory to slaughter can increase slightly, the rate of infection in such an area or location.

## 5. Conclusion

The result of this study is a clear pointer that haemoparasites are common with most animals slaughtered in abattoirs in Makurdi. A high prevalence of 24.6% was observed after parasitological analysis was done n the samples collected. One will therefore not be left in doubt that these animals carry this infection right from their places of production and as studies have revealed, the haemoparasites constitute great losses to farmers in terms of productivity, causing direct loss in milk yield, parturition rate, meat quantity and quality, and many other factors which may even lead to death of the animal if proper attention is not paid to these parasites.

## Data Availability

All data produced in the present study are available upon reasonable request to the authors

